# Impact of early life nutrition on gut health in children: A prospective clinical study

**DOI:** 10.1101/2021.02.19.21252039

**Authors:** Delphine Ley, Laurent Beghin, Jules Morcel, Florence Flamein, Charles Garabedian, Bertrand Accart, Elodie Drumez, Julien Labreuche, Frédéric Gottrand, Emmanuel Hermann

**Affiliations:** Univ. Lille, Inserm, CHU Lille, U1286 - INFINITE - Institute for Translational Research in Inflammation, F-59000 Lille, France; Univ. Lille, Inserm, CHU Lille, CIC-1403 Inserm-CHU, F-59000 Lille, France; Univ. Lille, CHU Lille, Department of Obstetrics & Gynecology, F-59000 Lille, France; Univ. Lille, CHU Lille, ULR 2694-METRICS: évaluation des technologies de santé et des pratiques médicales, F59000-Lille, France; CHU Lille - Inserm - UL, CRB/CIC1403, Lille, F-59037, France; CHU Lille, Department of Biostatistics, F59000-Lille, France

**Keywords:** Nutrition, breastfeeding, gut health, sIgA

## Abstract

**Introduction:** The first 1000 days of life could contribute to individual susceptibility to the later development of chronic non-communicable diseases. Nutrition in early life appears to be an important determinant factor for a sustainable child’s health. In this study, we propose to investigate the impact of exclusive breastfeeding on gut health in children.

**Methods and analysis:** A prospective cohort of newborns (n=350) will be recruited at birth and followed up to 4 years of age. The main objective is to evaluate the link between exclusive breastfeeding for at least 3 months and the gut health of the child at 4 years. The primary endpoint of assessment of gut health will be based on the non-invasive measurement of fecal secretory immunoglobulin A (sIgA) as a sensitive biomarker of the intestinal ecosystem. The presence of gastrointestinal disorders will be defined according to the clinical criteria of ROME IV. Information on parent’s nutritional habits and life style, breastfeeding duration and child’s complementary feeding will be collected along the follow-up. Cord blood cells and plasma at birth will be purified for further analysis. The meconium and stools collected at birth, 6 months, 2 years and 4 years of age will allow sIgA analysis.

**Ethics and dissemination:** This clinical study has obtained the approval from the national ethical committee. We plan to publish the results of the study in peer-review journals and by means of national and international conference.

**Trial registration number:** NCT04195425

**Strengths and limitations of the study:**

- This is a prospective and longitudinal mother/child cohort with minimal constraints (3 visits over 4 years) and minimal risk (no intervention and minimally invasive procedures).
- The study will collect a great deal of longitudinal information on children during the first years of life, their parents and their environment via questionnaires and biological samples (cord blood, meconium, stools).
- This study will obtain precise data on breastfeeding practices and their short- and medium-term effects on the health of the child, in particular and in an innovative way on gut health through stool samples for immunological analysis, and by using the ROME IV paediatric questionnaire.
- Weaknesses could be relied to confounding factors at the selection process and during the follow-up.
- Due to the non-interventional study design we will be able to study association but no causal relationship.
- The nutritional survey during the study is primarily based on reports from parents and is retrospective which could contribute to inaccuracies due to long delay between each visit.

## Introduction

The first 1000 days extending from conception to the end of the first two years of life define a period of intense development and growth of the fetus and infant. A growing body of evidence supports the central role of perinatal environment in programming individual predisposition to chronic non-communicable diseases ^1-5^. Many studies strongly suggest that perinatal factors such as mode of delivery, type of infant feeding or antibiotic therapy during the first months of life influence the later risk of chronic intestinal disease ^4 6^. However, the links between nutrition during the first 1000 days, gut development and lifelong health remain to be better understood.

Breastfeeding is the reference for infant feeding who should be exclusively breast-fed for at least the first 6 months for optimal growth ^7^. The influence of breastfeeding on short-term health and infant development has been demonstrated. In particular, breastfeeding is associated with superior cognitive development, decreased risk of ear, respiratory and gastrointestinal infections and atopic diseases ^8^. The consensus is much weaker with regard to longer-term beneficial effects of breastfeeding on outcome of chronic non-communicable diseases ^8-14^. Human milk contains all of the essential nutrients as well as bioactive factors that are thought to contribute to the health benefits of breastfed infants. Among these bioactive factors, secretory immunoglobulin A (sIgA), human milk oligosaccharides, lactoferrin, and milk fat globule membrane, have demonstrated role in shaping intestinal barrier function, immune development and composition of the intestinal microbiota, suggesting that breastmilk may have beneficial effect in programming long-term gut health ^5 6 15-17^. The understanding of imprint of early life nutrition on gut homeostasis is a concern for the development of preventive strategies aimed at promoting gut health.

We designed a prospective cohort PENSINE “Périnatalité, Environnement, Santé Intestinale et Nutrition de l’Enfant” to evaluate the impact of exclusive breastfeeding on gut health in children beyond 6 months of age and to identify the underlying mechanisms involved. The cohort of newborns (n=350) will be recruited at birth and followed up to 4 years of age. The nutritional habits and life style of parents, breastfeeding duration and complementary feeding of the child will be surveyed. Cord blood cells and meconium will be harvested at birth and stools will be collected at 6 months, 2 years and 4 years of age for implementing a biological collection. Gut health will be based on faecal sIgA known to regulate gut inflammation, intestinal microbiota richness and protection against gastro-intestinal infections ^18-20^. Immune analysis will be coupled to the evaluation of the presence of gastrointestinal disorders by using the ROME IV pediatric questionnaire.

## Study objective

### Primary objective

To evaluate the link between exclusive breastfeeding for at least 3 months and gut health at 4 years of age. Gut health will be evaluated based on the non-invasive measurement of faecal sIgA as a sensitive biomarker of the intestinal ecosystem ^21 22^. The presence of gastrointestinal disorders will be assessed using the ROME IV criteria ^23^. Stool samples collected from birth to 4 years of age will allow immunological studies.

### Secondary objectives

To evaluate the association between gut health (sIgA) at 4 years of age and the duration of exclusive breastfeeding.

To describe the mother’s eating habits during pregnancy (Food Frequency Questionnaire) and their links with gut health during the first 4 years of life.

To assess the association between the level of sIgA in stool during the first 4 years of life with the child’s growth and the presence of gastrointestinal disorders.

## Methods

### Study population and setting

Three hundred and fifty mother and newborn dyads will be included. The maternity enjoys around 5,600 births a year with a recruitment capacity fully satisfactory for our study. All pregnant women who intend to give birth at the Jeanne de Flandre maternity (Lille University Hospital) will be invited during the second trimester of pregnancy to participate in the PENSINE clinical study. Eligible pregnant women will be fully informed by the investigator with an information letter detailing the content and the relevance of the study. The inclusion of the pregnant women and future fathers will be consecutive and carried out during the third trimester of pregnancy after the signing of the informed consent by each of the parents. The inclusion of the child will take place after birth. Both parents will sign a second specific informed consent for the child’s participation.

The inclusion criteria will be as follows: parents ≥18 years old when signing the consent form; the ability to speak, read and write French, or being able to be assisted in completing questionnaires; pregnant mothers living in the Lille urban area; single pregnancy. Mothers who will be unable to participate in the study follow-up, i.e. mother under curatorship / guardianship or who participate in another clinical study incompatible with biological collection, will be excluded from the study. The exclusion criteria *a posteriori* for the child after birth will be: prematurity (<37 weeks of gestation), gastrointestinal congenital malformation, gastrointestinal surgery during the first six months of life and inability to participate in the study follow-up. Child will be also excluded from the study if they are under justice protection measure or if they participate in another clinical study incompatible with biological collection.

Pregnant mothers will be included in the study whether or not initiating breastfeeding. Exclusive breastfeeding will be defined as no intake of other liquids than breastfed (water, sugar water, herbal tea, fruit juice, pre- or pro-biotics supplementation, with the exception of oral rehydration solution or vitamins and medicines as stated by the World Health Organization). Among mothers who start breastfeeding at maternity, mixed breastfeeding will be considered when the child receives breast milk and infant formula, or other liquids. All children will be scheduled for three clinical visits, at 6 months, 2 years and 4 years of age.

The first inclusions have been done in December 2020. The study will last 5 years. All administrative and regulatory issues will be managed by the Lille University Hospital Health Research Directorate including data protection and ethical. The logistical coordination of this project will be under the responsibility of the paediatric Clinical Investigation Center (CIC-1403-Inserm-CHU) of the Lille University Hospital involved in clinical data handling and biological samples collection.

### Study design

It will be a prospective clinical study including 350 mother / child dyads recruited at the Lille University Jeanne de Flandre maternity (Figure 1). The follow-up of the children will be carried out from birth until the age of 4 years. The diagram of the research process is divided into three periods (Figure 2): prenatal period (period 1: (inclusion of the pregnant mother, study of the nutritional profile and lifestyle of the pregnant woman), birth of the child (period 2: inclusion of the child and collection of perinatal factors), followed by 4 years (period 3: clinical and nutritional follow-up of the child).

**Figure 1.**
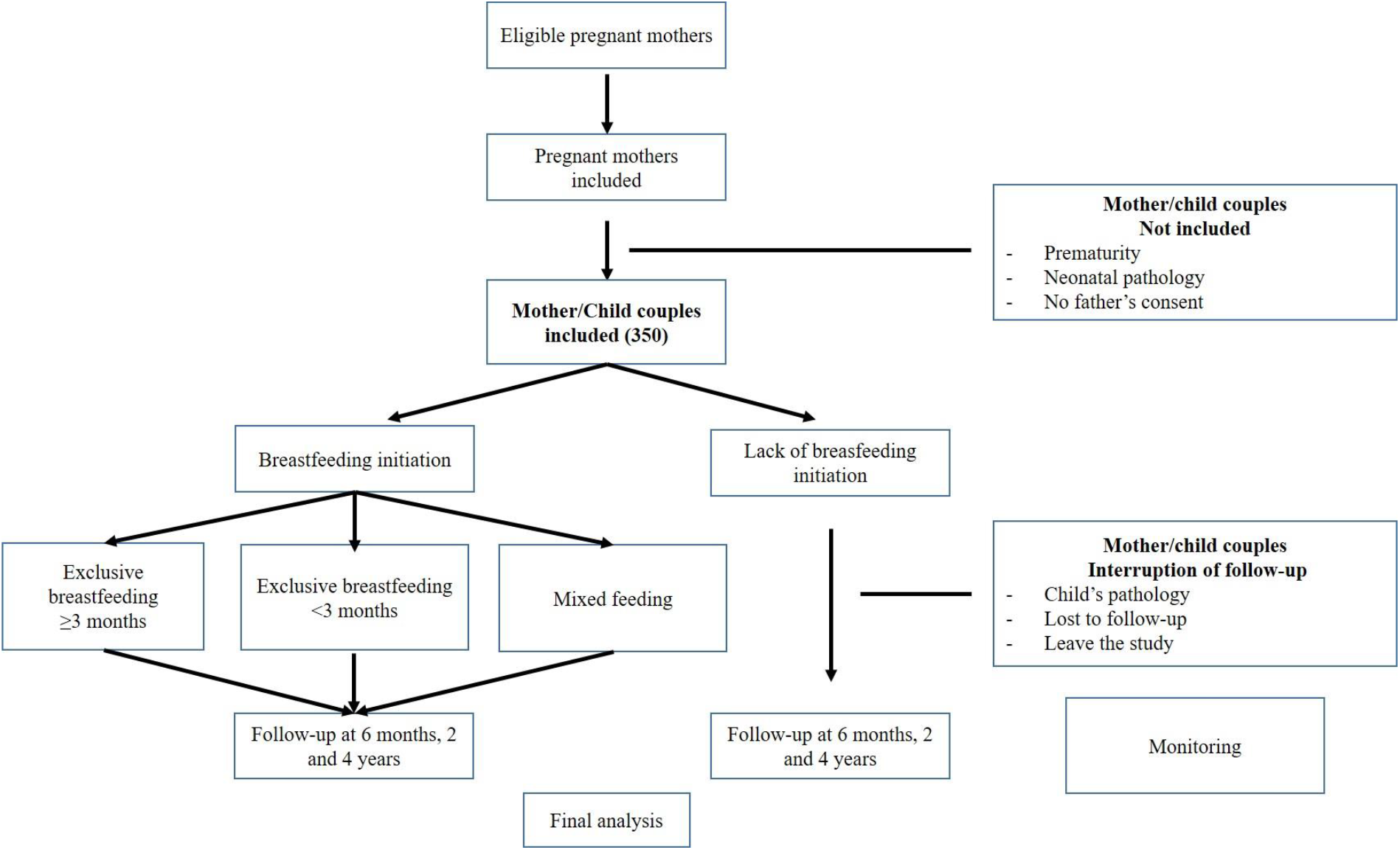
Study design

**Figure 2.**
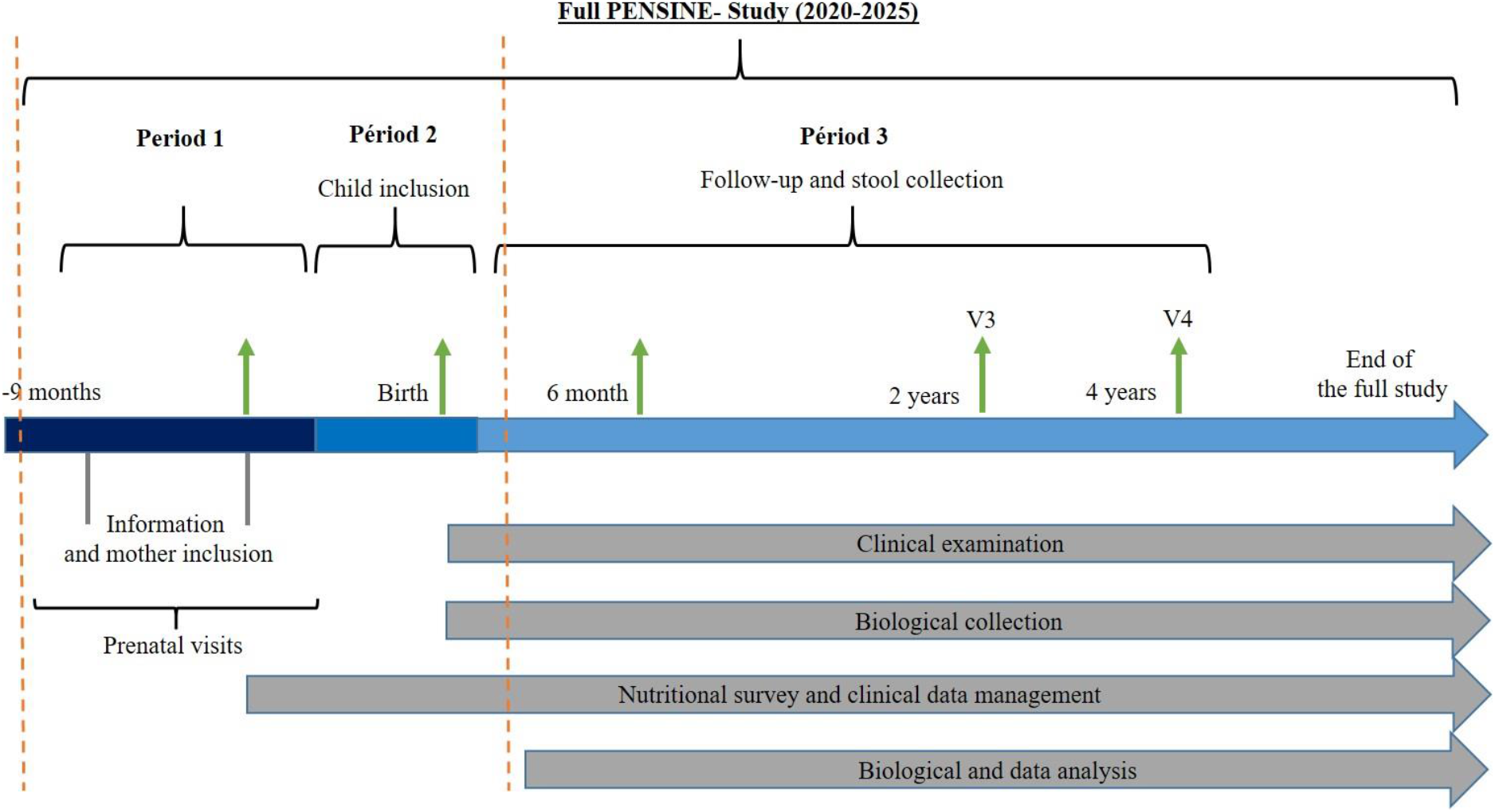
Study schedule

i. Prenatal period (1): During the third trimester of pregnancy, mothers will have to filled out a questionnaire on their socio-economic level and a QFA questionnaire (Questions of Food Frequency) relating to their eating habits, providing information on their consumption of meat and fish, fruit or vegetables, dairy products, fat or sugar, industrial products. Health information of the mother and medication received including antibiotics during pregnancy, as well as gastrointestinal history of parents, will be collected from the obstetric medical record.
ii. Birth period (2): Birth information, including mode of delivery and per partum antibiotic, will be collected from the mother’s obstetrical record. Cord blood will be taken to store peripheral blood mononuclear cells (CBMC) and fetal plasma as part of a biological bank. Children’s meconium will be collected to be frozen and stored.
iii. Follow-up Period (3) after birth: Three follow-up visits will be scheduled at 6 ± 1 month, 24 ± 1 months, and 48 ± 1 months at the pediatric CIC. The visit will include: clinical examination of the child, anthropometric measurements, collection of medical information including episodes of fever, diarrhea or atopic dermatitis and medication received, standardized food questionnaires (Food and life style question, see table 1), and questionnaires intended to diagnose the presence of gastrointestinal disorders in children (ROME IV). The stools of infants will be collected at each time, aliquot and stored for further immunological studies (faecal sIgA and inflammatory markers such as faecal calprotectin) and gut microbiota analysis.

**Table 1.**
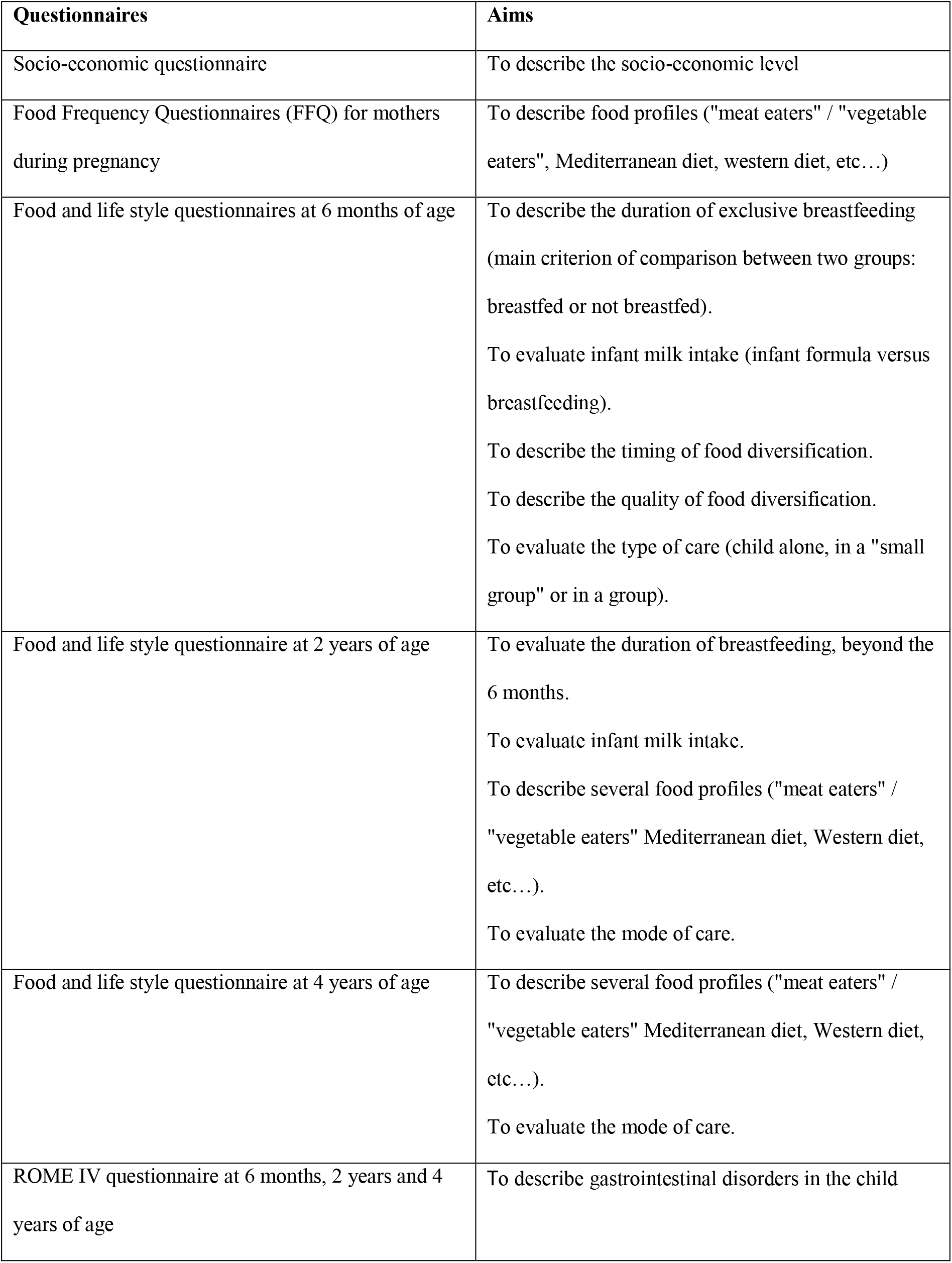
Objectives of PENSINE surveys

### Sample size

The study was powered to demonstrate an effect size (standardized mean difference) in sIgA at 4-years ≥0.40 considered as medium. This effect size was chosen given the effect size of exclusive breastfeeding on gut health measured by sIgA at 3 months of life reported in literature^10^. To demonstrate this effect size at 4 years, assuming an exclusive breastfeeding rate of 40% at 3-month of age, with a two-sided test at 5% of significance and statistical power of 80%, 208 subjects (mother / child dyad) are needed. To take into account 40% of subjects who cannot be analyzed at 4 years, 350 subjects should be included. Because premature infants are excluded, the inclusion of pregnant mothers in the 3rd trimester of pregnancy will take place until the goal of the inclusion of 350 eligible full-term newborns at birth is reached. Considering 10% prematurity, we anticipate the inclusion of 390 pregnant mothers.

### Data and biobanking management

The study will provide a set of medical and dietary outcomes for each participant during pregnancy, at birth, and during the follow-up. The individual data collected during the study will be reported on a source document and then will be recorded into a computer database (electronic-case report form (e-CRF)). The processing of data will be carried out under the conditions of confidentiality defined by the French law of January 6, 1978 modified relating to data processing, files and freedoms (Commission Nationale de l’Informatique et des Libertés (CNIL)) as well as in accordance with European Regulation (General Data Protection Regulation: RGPD). Access to the data will be restricted to those directly involved in the study. Statistical analysis will be performed by the Methodology, Biostatistics and Data Management Department at the Lille University Hospital. The data concerning this study will be archived for a minimum period of fifteen years from the end of the research or its early termination without prejudice to the legislative and regulatory provisions in force.

### Medical and anthropometric data

Medical data will be recorded at baseline by trained staff will include complications during pregnancy and/or childbirth, mode of delivery, birth weight, length and head circumference of the new-born. Anthropometric parameters will be measured as part of clinical assessment at birth, 6 months, 2 years and 4 years of age in infants and children with standardized techniques. Information on the exposure of relevant drugs will be recorded at baseline and during the study (using standardized questionnaires). The presence of gastrointestinal disorders will be assessed by a pediatric gastroenterologist using the pediatric ROME IV diagnostic criteria ^24^ and recorded in the e-CRF.

### Biobanking

According to the main objective, an aliquot of stool from 4-year-old children will be used to measure sIgA by ELISA (Enzyme-Linked Immunosorbent assay) methodology (commercial kit). The other biological elements of the biobank (cord blood plasma, CBMC, meconium, aliquots of stool) will be available as part of ancillary research projects by our team or open to the scientific and clinical community. These research projects might include analysis of immunological biomarkers, composition of the intestinal microbiota on stool samples and epigenetics on CBMC.

All samples will be kept frozen at the Biological Resource Centre (BRC) of the Lille University Hospital to constitute a biobank until the analysis. The BRC/CIC will ensure the management of biological samples with a view to their subsequent analysis, storage and management of these in its aspects of referencing, storage, traceability of inputs, outputs or incidents.

### Blood samples

After delivery, cord blood will be drawn in heparinized tubes by the midwife from the umbilical vein after clamping and before cutting the cord. The labeled tube will be transferred to the BRC for processing. Plasma and purified CBMCs will be frozen and stored as part of the protocol. After collecting and distributing the plasma in an aliquot, CBMCs will be purified and cryopreserved in liquid nitrogen for biobanking.

At the age of 4 years, a finger prick will be proposed to collect a drop of blood of the child using a lancing device. The drop of blood will be placed on a Whatman filter paper using the dried blood spot technique for subsequent analysis of inflammatory serological biomarkers.

### Fecal samples

At birth, meconium will be collected under sterile conditions, transferred to the BRC for aliquoting and freezing. At 6 months, 24 months, and 48 months the child’s stool will be collected by the parents at home in the days before the visit and then placed in a labeled sterile container before storing at +4°C. The stool sample will be brought back to the CIC for the follow-up visit. The sample will be immediately transferred to the BRC in order to aliquot, manage traceability and storage at −80°C for further analysis.

### Statistical analysis

Statistical analysis will be independently performed by the Biostatistics Department of the Lille University Hospital under the responsibility of Julien Labreuche and Elodie Drumez. Data will be analyzed using the SAS® software (SAS® Institute Inc, Cary, NC, USA) and all statistical tests will be performed with a 2-tailed alpha risk of 0.05; all secondary objectives will be considered as exploratory and no correction for multiple comparisons will be applied. A detailed statistical analysis plan will be written and finalized prior to the database lock.

Primary objective: the sIgA level assessed at 4-year of life will be compared according to exclusive breastfeeding status at 3-month of age using a Student t test; the standardized mean difference will be calculated with its 95% confidence interval. To account pre-specified confounding factors (age of dietary diversification, socio-economic level of parents, gestational age at birth, mode of delivery, antibiotic therapy during the first months), a multivariable linear regression model will be used, with sIgA level as dependent variable, and exclusive breastfeeding status at 3-month of age, and pre-specified confounding factors as independent variables. To account missing data on dependent and independent variables, primary analysis will be performed after handling missing values by multiple imputation procedure under the missing at random assumption using a regression switching approach (chained equation with number of imputation (m) guided by maximal fraction of missing information (FMI)/m<0.1) ^25^. The imputation models will be used all baseline and 3-month characteristics, and study outcome (sIgA level at 4-year of life) with predictive mean matching method for quantitative variables and logistic regression (binary, ordinal, or polynomial) for categorical variables). Estimates obtained in the different imputed data sets will be combined using Rubin’s rules ^26^. A complete-case analysis will be done as first sensitivity analysis. A second sensitivity analysis will be done by using the Hayden method to account the rate of missing information on primary outcome, by using a weighted linear regression model ^27^. The weights will be calculated as the probabilities of missing outcome value in each group of interest estimated using multivariable logistic regression model including missing information status as dependent variable and all baseline and 3-month characteristics as independent variables.

Secondary objectives: Briefly, to evaluate the association of sIgA at 4 years of age with the duration of exclusive breastfeeding (assessed as ordinal 5-level categorical variable) a one-way analysis of variance will be performed; a trend test will be done using linear contrast. To account pre-specified confounding factors, a multivariable linear regression model will be performed. To assess the association of mother’s eating habits during pregnancy (measured by Food Frequency Questionnaire) and the gut health during the first 4 years of life (assessed at 6-month, 2- and 4-year), we will used a linear mixed model (with unstructured covariance matrix to account the repeated measures), by included mother’s eating habit characteristics, time and time* mother’s eating habit characteristics as fixed effects. Post-hoc comparison at each time of follow-up will be done using linear contrast. As specified above, multivariate analysis will be done by considering the same pre-specified confounding factors into linear mixed model. We also used the same approach to assess the association of child’s growth and the presence of gastrointestinal disorders and the level of sIgA in stool first 4 years of life (assessed at 6-month, 2- and 4-year). For all linear regression models, the normality of residuals will be checked.

### Perspectives

The study will allow the child’s health trajectory to be understood as a function of gut health and the complex intricacies between multiple factors including the perinatal environmental exposure, nutritional status, inflammation, microbiota, or epigenetic modifications. The data from the complete study will provide a conceptual basis for thinking about the possibility of nutritional recommendations for mothers and children with regard to a sustainable health trajectory. The biobank will also open up transversal perspectives for clinical research with a significant impact on clinicians in the field of neonatology, obstetrics, pediatrics and nutrition.

### Ethics and dissemination

The clinical study was approved by the French national ethical committee (reference CPP / AU 1547/ 18^th^ October 2019). It will be conducted in accordance with the clinical study protocol, and with the Good Clinical Practices. In particular, the trial participant (legal representative of the child) will receive fair and complete information through the information letter, explained orally by the investigator or the midwife. Each legal representative of the child will give their written consent on a consent form prior to fill out the questionnaires and to collect biological samples. The study is registered in the public ClinicalTrials.gov database (NCT04195425).

The first deliverables of the study will consist in establishing regular reports, and at the end of the study a final report will be written by the medical investigator. This vast study will lead to clinical and biological results that will be communicated through publications and communications by poster or oral at specialist congresses or seminars. Our study is totally in the field of early health programming, which is the main theme of the DOHAD (Developmental Origins of Health and Disease), which organizes conferences in which many elements of our study might be presented. In conclusion, the identification of risk profiles for intestinal diseases and the rapid introduction of new recommendations to improve health outcomes will be major outlets for this study. Collectively, the overall project will produce appropriate knowledge useful for regulatory and risk management agencies and even for related stakeholders in order to support specific recommendations.

## Data Availability

The individual data collected during the study will be reported on a source document and then will be recorded into a computer database (electronic-case report form (e-CRF)). The processing of data will be carried out under the conditions of confidentiality defined by the French law of January 6, 1978 modified relating to data processing, files and freedoms (Commission Nationale de l Informatique et des Libertes (CNIL)) as well as in accordance with European Regulation (General Data Protection Regulation: RGPD). Access to the data will be restricted to those directly involved in the study.

## Acknowledgments

We thanks all people who contributed in the development of the protocol. We thank the biological resource centre for biobanking

## Authors’ contributions

DL, LB and EH designed the project and drafted the manuscript. FG supervised the project. FL, CG, BA, ED, JL reviewed the manuscript. All authors read and approved the final version of the manuscript.

